# Comparative effectiveness of four COVID-19 vaccines, BNT162b2 mRNA, mRNA-1273, ChAdOx1 nCov-19 and NVX-CoV2373 against SARS-CoV-2 B.1.1.529 (Omicron) infection

**DOI:** 10.1101/2022.12.22.22283869

**Authors:** Bette Liu, Sandrine Stepien, Jiahui Qian, Heather Gidding, Katrina Nicolopoulos, Janaki Amin, Allen Cheng, Kristine Macartney

## Abstract

There is limited data directly comparing the effectiveness of COVID-19 vaccines. We compared rates of SARS-CoV-2 Omicron BA.1/2 infection in Australian adults during March to May 2022 who had received one of four COVID-19 vaccines in the last 14-63 days as either a primary course or a booster dose. As a primary course, compared to recipients of BNT162b2 mRNA vaccine, adjusted hazard ratios for SARS-CoV-2 infection were 1.03 (95%CI 0.82-1.30), 1.19 (0.95-1.49), 1.70 (1.46-1.97) for respectively mRNA-1273, ChAdOx-1 nCov-19 and NVX-CoV2373. For booster dose, respective adjusted hazard ratios compared to BNT162b2 mRNA vaccine were 1.02 (95%CI 1.00-1.04), 1.20 (1.10-1.32), 1.39 (1.20-1.60). Our findings suggest relatively higher effectiveness of ancestral strain mRNA vaccines against SARS-CoV-2 Omicron infection than viral vector and protein subunit vaccines, but further studies are required due to small numbers of recipients of ChAdOx-1 nCov-19 and NVX-CoV2373.

## Background

There are currently 11 COVID-19 vaccines granted emergency use listing (EUL) by the World Health Organization (WHO)^1^. While all have been shown to be efficacious in controlled clinical trials in a primary course, data comparing effectiveness between vaccine types as a primary course and booster are limited. Direct comparisons between vaccine types are difficult because licensure and vaccine introduction into national programs has occurred at different time points during the SARS-CoV-2 pandemic; dominant virus variants and underlying population-level infection-induced immunity also differ. Additionally, clinical recommendations regarding use by vaccine type, such as the eligible population for primary and booster dosing, and intervals between doses have changed.

Four vaccines, BNT162b2 mRNA (Pfizer Comirnaty), mRNA-1273 (Moderna Spikevax), ChAdOx1 nCov-19 (Oxford-Astrazeneca Vaxzevria) and NVX-CoV2373 (Novavax Nuvaxovid) are available for use in Australian adults^2^. BNT162b2 mRNA and ChAdOx1 nCov-19 were available in early 2021; most of the Australian adult population received these vaccines in a homologous 2 dose primary course^3^. mRNA-1273 was available from October 2021 and NVX-CoV2373 from February 2022, meaning that substantially fewer individuals received these latter two vaccines as a primary course. Booster doses (third dose) of COVID-19 vaccines were recommended from late November 2021 in response to evidence of waning protection from primary vaccination and significant community transmission of the SARS-CoV-2 Omicron variant in Australia.

To address the limited data on comparative effectiveness of COVID-19 vaccine types, particularly for NVX-CoV2373, we sought to directly compare effectiveness of the four vaccines currently used in Australia against infection with SARS-CoV-2 during the period March-May 2022 when the Omicron BA.1 and BA.2 variants were circulating.

## Methods

### Study design, data sources, definitions

We constructed a cohort of individuals aged ≥18 years residing in the areas of Greater Sydney and Hunter New England in Australia’s most populous state, New South Wales (NSW). The cohort was defined using the Australian Immunisation Register (AIR), a national registry including all people in Australia registered with Australia’s national universal health insurance scheme (Medicare) as well as individuals not registered but who had received a COVID-19 vaccine. The AIR includes demographic information on individuals and vaccine details (brand, date of administration, dose number). Reporting of COVID-19 vaccination to the AIR was made mandatory prior to the COVID-19 vaccine program commencement.

The cohort was probabilistically linked (using name, date of birth, residential address and sex) to notifications of SARS-CoV-2 infections and death registrations. Notification of SARS-CoV-2 infection was mandatory to record in the NSW Notifiable Conditions Information Management System (NCIMS) with diagnoses made by polymerase chain reaction (PCR) tests or rapid antigen tests (RAT) included during the study period. Death registrations included all registered deaths in NSW and the date of death. The main study outcome was a SARS-CoV-2 infection, as based on NCIMS case notification. Vaccination data were obtained from each individual’s AIR record. Our analyses compared vaccine type for the primary course and the booster. For the primary course, we classified individuals as recipients of one of the four vaccine types if both dose 1 and 2 were the same vaccine type, heterologous primary course recipients were categorised separately. For the booster dose comparison we compared the vaccine type received for dose 3, regardless of the primary course type.

### Statistical analysis

We conducted separate analyses for the primary course and for boosters. The study period commenced on 1 March 2022 and ended on 27 May 2022, a period corresponding to when the Omicron subvariants BA.1 and BA.2 were circulating^4^. For the main analyses we excluded individuals who prior to the study period had a COVID-19 case notification, death record, or had received more vaccine doses than were of interest for the analysis (dose 3 for the primary course analysis and dose 4 for the booster analysis). We used Cox-regression to estimate hazard ratios comparing infection rates between recipients of different vaccine types. Vaccination status, including type, dose and interval since receipt was treated as a time-varying variable. Analysis time commenced on 1 March 2022 and ended at either the date of COVID-19 notification, death, receipt of a 3^rd^ dose (for the primary course analysis) or 4^th^ dose (for the booster dose analysis) or the 27 May 2022 which was the last date of complete records.

To ensure comparability of the interval since receipt of vaccine, comparisons focused on the interval 14-63 days (2-6 weeks) after receipt of the second dose for the primary course analysis, and 14-63 days after receipt of the 3^rd^ dose for the booster analysis. The BNT162b2 mRNA (Pfizer Comirnaty) vaccine was set as the reference group as this was the most widely used vaccine type for both primary and booster doses. Analyses were adjusted for age (in 2-year increments), gender, socioeconomic status (deciles based on an Australian index using residence and census data^5^), and number of comorbidities (based on ICD-10 coded hospitalisations with specified medical conditions in the 2 years prior to the analysis start date^3^).

We conducted subgroup analyses by age (<50 and ≥50 years), gender and socioeconomic status and also conducted two sensitivity analysis; 1) including individuals who had had a prior SARS-CoV-2 case notification and adjusting for prior infection and; 2) restricting analyses to the period from 1 April to 27 May 2022 when more individuals had had an opportunity to have NVX-CoV2373. This study was approved by the NSW Population and Health Services Research Ethics Committee (2022/ETH00584).

## Results

After exclusions, the main study cohort for the primary course analysis included 2.27 million individuals and the booster analysis 4.61 million. However, for the main comparisons, of those who received 2 or 3 doses within 14-63 days, the number of individuals classified by the end of follow up was 5923 and 204748, respectively. Table 1 and 2 show characteristics of the cohort overall and by vaccine type as classified at the end of follow-up noting that because of the time-varying nature of the exposure these groups do not correspond precisely to those for which the hazard ratios were estimated but are shown to provide an indication of the differences in those receiving each vaccine type during the observation period. The mean age in the primary course analysis was 44.0 years (standard deviation [SD] 17.6 years) and 47.7% were female; for the booster dose the mean age was 48.5 (SD 18.5 years) and 50.4% were women.

**Table 1:** Characteristics of cohort for primary course analyses overall and according to vaccine type received within 14-63 days of second dose at end of follow-up

|  | Primary course vaccine type, second dose received within 14-63 days |  |  |  |  |  |  |  |  |  |
| --- | --- | --- | --- | --- | --- | --- | --- | --- | --- | --- |
|  | BNT162b2 mRNA |  | mRNA-1273 |  | ChAdOx1 nCov-19 |  | NVX-CoV2373 |  | Total population |  |
|  | n | % | n | % | n | % | n | % | n | % |
| **N** | 3212 |  | 340 |  | 289 |  | 2082 |  | 2273324 |  |
| **Mean age in years (SD)** | 42.6 (19.0) |  | 42.6 (17.8) |  | 44.0 (17.7) |  | 46.5 (16.7) |  | 44.0 (17.6) |  |
| **Age, years** |  |  |  |  |  |  |  |  |  |  |
| 18-29 | 951 | 29.61 | 96 | 28.24 | 68 | 23.53 | 340 | 16.33 | 555853 | 24.45 |
| 30-39 | 849 | 26.43 | 77 | 22.65 | 68 | 23.53 | 499 | 23.97 | 523069 | 23.01 |
| 40-49 | 449 | 13.98 | 62 | 18.24 | 62 | 21.45 | 428 | 20.56 | 395597 | 17.40 |
| 50-59 | 311 | 9.68 | 43 | 12.65 | 33 | 11.42 | 306 | 14.70 | 329639 | 14.50 |
| 60-69 | 287 | 8.94 | 27 | 7.94 | 22 | 7.61 | 289 | 13.88 | 246316 | 10.84 |
| 70-79 | 169 | 5.26 | 23 | 6.76 | 22 | 7.61 | 157 | 7.54 | 131847 | 5.80 |
| >80 | 196 | 6.10 | 12 | 3.53 | 14 | 4.84 | 63 | 3.03 | 91003 | 4.00 |
| **Socioeconomic status in quintiles** |  |  |  |  |  |  |  |  |  |  |
| 1 (most disadvantaged) | 1065 | 33.16 | 101 | 29.71 | 64 | 22.14 | 376 | 18.06 | 508625 | 22.37 |
| 2 | 668 | 20.79 | 80 | 23.53 | 65 | 22.49 | 407 | 19.55 | 478236 | 21.19 |
| 3 | 659 | 20.51 | 59 | 17.35 | 60 | 20.76 | 454 | 21.81 | 462300 | 20.34 |
| 4 | 434 | 13.51 | 57 | 16.77 | 59 | 20.41 | 433 | 20.79 | 428301 | 18.84 |
| 5 (most advantaged) | 386 | 12.02 | 43 | 12.65 | 41 | 14.19 | 412 | 19.78 | 395862 | 17.41 |
| **Gender** |  |  |  |  |  |  |  |  |  |  |
| Female | 1626 | 50.62 | 174 | 51.18 | 121 | 41.87 | 1093 | 52.50 | 1083347 | 47.65 |
| Male | 1586 | 49.38 | 166 | 48.82 | 168 | 58.13 | 989 | 47.50 | 1189977 | 52.35 |
| **Co-morbidities** |  |  |  |  |  |  |  |  |  |  |
| No | 2931 | 91.25 | 313 | 92.06 | 265 | 91.70 | 1983 | 95.24 | 2186387 | 96.18 |
| Yes | 281 | 8.75 | 27 | 7.94 | 24 | 8.30 | 99 | 4.76 | 86937 | 3.82 |
| **Median interval between dose 1 and 2 in days (IQR)** | 35 (22-121) |  | 35 (28-84) |  | 81 (35-161) |  | 24 (21-29) |  |  |  |

**Table 2:** Characteristics of cohort for booster analyses overall and according to vaccine type received within 14-63 days of third dose at end of follow-up

|  | Booster vaccine type, third dose received within 14-63 days |  |  |  |  |  |  |  |  |  |
| --- | --- | --- | --- | --- | --- | --- | --- | --- | --- | --- |
|  | BNT162b2 mRNA |  | mRNA-1273 |  | ChAdOx1 nCov-19 |  | NVX-CoV2373 |  | Total population |  |
|  | n | % | n | % | n | % | n | % | n | % |
| **N** | 160636 |  | 40876 |  | 1351 |  | 1885 |  | 4610877 |  |
| **Mean age in years (SD)** | 43.8 (17.2) |  | 42.9 (16.7) |  | 54.8 (17.8) |  | 52.9 (16.6) |  | 48.5 (18.5) |  |
| **Age, years** |  |  |  |  |  |  |  |  |  |  |
| 18-29 | 37119 | 23.11 | 10317 | 25.24 | 147 | 10.88 | 183 | 9.71 | 832275 | 18.05 |
| 30-39 | 39598 | 24.65 | 9991 | 24.44 | 199 | 14.73 | 303 | 16.07 | 883918 | 19.17 |
| 40-49 | 29888 | 18.61 | 6926 | 16.94 | 155 | 11.47 | 292 | 15.49 | 794592 | 17.23 |
| 50-59 | 22019 | 13.71 | 5896 | 14.42 | 220 | 16.28 | 368 | 19.52 | 742232 | 16.10 |
| 60-69 | 16934 | 10.54 | 4788 | 11.71 | 319 | 23.61 | 418 | 22.18 | 644538 | 13.98 |
| 70-79 | 9435 | 5.87 | 2003 | 4.90 | 221 | 16.36 | 244 | 12.94 | 444348 | 9.64 |
| >80 | 5643 | 3.51 | 955 | 2.34 | 90 | 6.66 | 77 | 4.08 | 268974 | 5.83 |
| **Socioeconomic status in quintiles** |  |  |  |  |  |  |  |  |  |  |
| 1 (most disadvantaged) | 30442 | 18.95 | 6580 | 16.10 | 301 | 22.28 | 224 | 11.88 | 877469 | 19.03 |
| 2 | 33725 | 20.99 | 9035 | 22.10 | 291 | 21.54 | 305 | 16.18 | 922816 | 20.01 |
| 3 | 33884 | 21.09 | 8239 | 20.16 | 232 | 17.17 | 382 | 20.27 | 931620 | 20.20 |
| 4 | 32643 | 20.32 | 8249 | 20.18 | 269 | 19.91 | 430 | 22.81 | 932625 | 20.23 |
| 5 (most advantaged) | 29942 | 18.64 | 8773 | 21.46 | 258 | 19.10 | 544 | 28.86 | 946347 | 20.52 |
| **Gender** |  |  |  |  |  |  |  |  |  |  |
| Female | 87297 | 54.34 | 21928 | 53.65 | 671 | 49.67 | 1139 | 60.42 | 2323770 | 50.40 |
| Male | 73339 | 45.66 | 18948 | 46.35 | 680 | 50.33 | 746 | 39.58 | 2287107 | 49.60 |
| **Co-morbidities** |  |  |  |  |  |  |  |  |  |  |
| No | 149944 | 93.34 | 38804 | 94.93 | 1180 | 87.34 | 1728 | 91.67 | 4307387 | 93.42 |
| Yes | 10692 | 6.66 | 2072 | 5.07 | 171 | 12.66 | 157 | 8.33 | 303490 | 6.58 |
| **Primary course vaccine type** |  |  |  |  |  |  |  |  |  |  |
| BNT162b2 mRNA | 107872 | 67.15 | 22378 | 54.75 | 61 | 4.52 | 1084 | 57.51 |  |  |
| ChAdOx1 nCov-19 | 50881 | 31.67 | 16041 | 39.24 | 1264 | 93.56 | 715 | 37.93 |  |  |
| Other | 1883 | 1.17 | 2457 | 6.01 | 26 | 1.92 | 86 | 4.56 |  |  |

Of the 5923 who received the second dose of their primary course in the last 14-63 days by end of follow-up, BNT162b2 mRNA was mostly commonly received (N=3212) followed by NVX-CoV2373 (n=2082). NVX-CoV2373 recipients were older, more likely female, of higher socioeconomic status, but less likely to have a comorbidity than mRNA vaccine recipients. ChAdOx1 nCov-19 recipients were older, and less likely female than mRNA vaccine recipients. The interval between receipt of first and second doses of NVX-CoV2373 were shorter (median 24 days) than for mRNA vaccine recipients (median 35 days) and ChAdOx1 nCov-19 recipients (median 81 days).

Among the 204748 who were within 14-63 days of receipt of their first booster (3^rd^ dose), the majority received an mRNA vaccine (BNT162b2=160363 (78%); mRNA-1273=40876 (20%)) with <1% receiving either of NVX-CoV2373 or ChAdOx1 nCov-19. Similar to the primary course, NVX-CoV2373 and ChAdOx1 nCov-19 booster recipients were older than mRNA booster recipients. NVX-CoV2373 recipients were also more likely to be women and were less likely socioeconomically disadvantaged than mRNA booster recipients. Most individuals had received BNT162b2 for their primary vaccine course although this differed by the booster vaccine type; 67.2% of BNT162b2 booster recipients also had a BNT162b2 primary course while only 4.5% of ChAdOx1 nCov-19 booster recipients had BNT162b2 as the primary course.

Figure 1 shows case numbers, crude rates, and adjusted hazard ratios (aHR) for SARS-CoV-2 infection by vaccine type of primary course or booster. The crude infection rate was slightly higher in booster dose recipients compared to primary course recipients. Among recent BNT162b2 recipients diagnosed with COVID-19, 49% (primary) and 50% (booster) were diagnosed using PCR; this was higher among recent NVX-CoV2373 recipients (53% for primary; 58% for booster). The relative SARS-CoV-2 infection rate by vaccine type was similar for both primary course and booster; similar infection rates were evident among mRNA vaccine recipients (comparing mRNA-1273 to BNT162b2, primary course aHR was 1.03 (95%CI 0.82-1.30); booster aHR 1.02 (95%CI 1.00-1.04)). Recipients of ChAdOx1 nCov-19 for both the primary and booster had slightly higher infection rates compared with BNT162b2, although not significantly different for the primary course (primary course aHR 1.19 (95%CI 0.95-1.49); booster aHR 1.20 (95%CI 1.10-1.39)). For NVX-CoV2373, the infection rates were higher than for BNT162b2 for both primary (aHR 1.70 (95%CI 1.46-1.97) and booster doses (aHR 1.39 (95%CI 1.20-1.60)). These infection rate differences by vaccine type were generally consistent in subgroups of individuals <50 and 50+ years of age, in men and women, and by lower or higher socioeconomic status (Supplementary Table 1).

**Figure 1:**
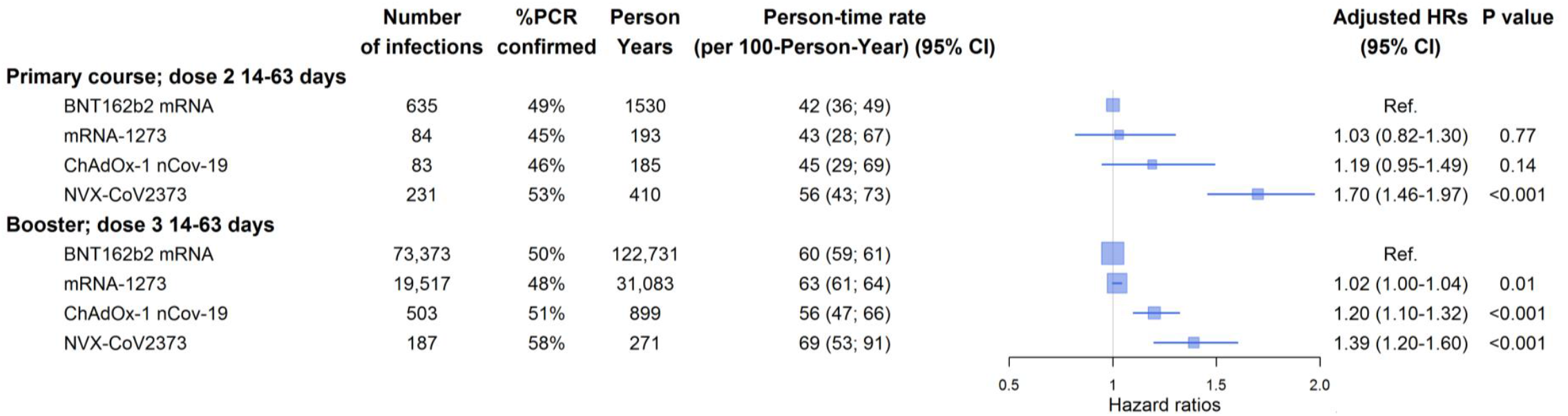
Rate and hazard ratio for SARS-CoV-2 infection by primary course and booster vaccine type in interval 14-63 days following receipt of dose, March-May 2022. Hazard ratios adjusted for age, gender, socioeconomic status and comorbidity. Boxes in plot are proportional to the amount of data and lines represent the 95% confidence intervals of the hazard ratio. Analyses included only individuals without a prior COVID-19 diagnosis. Total study population included in primary course analysis N=2,273,324; booster analysis N=4,610,877.

Sensitivity analyses shown in Supplementary Table 2 included an additional 424,033 (primary course) and 637,476 (booster) individuals who had previously been notified with SARS-CoV-2 infection. When prior infection was adjusted for, results were compatible with the main analyses. Similarly, when only the period from 1 April to 27 May was examined (Supplementary Table 3), while there were smaller numbers of individuals and events, results were also consistent.

## Discussion

This study directly compares four of the WHO EUL COVID-19 vaccines, representing 3 different vaccine platforms, in the same calendar period of the SARS-CoV-2 pandemic, when the BA1 and BA.2 Omicron subvariants were circulating. We found, compared to the BNT162b2 mRNA vaccine, that mRNA-1273 had similar effectiveness against SARS-CoV-2 infection when used either as a primary course or as a booster. Adjusted hazard ratios for COVID-19 in recipients of the adenoviral vector vaccine, ChAdOx1 nCov-19, and protein subunit vaccine NVX-CoV2373 were higher than in those who received the mRNA vaccines when used either as a primary course or as a booster.

There are few studies comparing effectiveness of COVID-19 vaccine types, particularly protein subunit vaccines. A recent systematic review and meta-analysis compared vaccine efficacy or effectiveness (from trials and observational studies) between vaccine platform types (mRNA, viral vector, inactivated and protein subunit) against variants of concern prior to Omicron (Alpha, Beta, Gamma, Delta) for primary and booster vaccination and reported that VE for primary vaccination was higher with mRNA vaccines compared to all other types but did not separate the other vaccine platform types due to smaller numbers of studies^6^. Most studies examining effectiveness against the Omicron variant have focussed on mRNA vaccines given as a booster with studies suggesting similar effectiveness of the BNT162b2 and mRNA-1273 vaccines against severe disease^7,8^, regardless of the priming course^7^. However, one study suggested that mRNA-1273 elicited higher protection against infection with the Omicron variant when compared to BNT162b2 in individuals primed with BNT162b2^9^. Our findings are mostly consistent with the meta-analyses and studies comparing the two mRNA vaccines and ChAdOx-1. However, this is the first report that directly compares effectiveness of the protein subunit vaccine NVX-CoV2373 to other COVID-19 vaccine types.

Our findings on comparative effectiveness by type of booster are somewhat consistent with published data from the COV-Boost randomised trial^10^ which compared immunogenicity to ancestral spike protein between seven vaccine types including all four vaccine types examined here, given as a booster in individuals primed with either ChAdOx1 nCov-19 or BNT162b2. COV-Boost found the highest spike IgG GMCs in those receiving mRNA-1273 followed by BNT162b2, NVX-CoV2373 and then ChAdOx1 nCov-19, noting that the mRNA-1273 dose used in the trial was double that registered and used in Australia (100µg vs 50µg). The relative neutralising antibody titres in this trial were also similar in individuals primed with ChAdOx1 nCov-19. Another study which compared neutralising activity of several COVID-19 vaccines against different Omicron sublineages found greater fold decreases of neutralisation against both BA.1 and BA.2 sublineages for NVX-CoV2373 compared to BNT162b2 and mRNA-1273^11^. A recent predictive model^12^ suggested that vaccine protection against SARS-CoV-2 infection decreases with increasing genetic distance from the original vaccine-type strain, but that the level of decrease differs according to vaccine technology platform; reduced protection from protein subunit vaccines compared to mRNA and viral vector vaccines was predicted. Our study results during a period of circulation of Omicron BA.1 and BA.2 strains, highly divergent from the ancestral virus, are consistent with this model’s findings.

It is important to note that we only examined infection and did not have information to confirm if this resulted in clinical disease. We lacked data on severe disease but, it is unlikely the relatively small numbers of individuals receiving primary or booster doses of NVX-CoV2373 or ChAdOx1 nCov-19 during the study period would have allowed sufficient statistical power to compare effectiveness across the 4 vaccine types for this outcome. Uncontrolled confounding by behavioural characteristics, such as differences in propensity to test, among those who preferred non-Mrna vaccine types cannot be excluded as a possible explanation for the differences we found. However, our results were consistent in subgroups of age, socioeconomic status and gender as well as in various sensitivity analyses. We also noted that while there were differences in the interval between receipt of dose 1 and 2 in the primary course analysis with recipients of NVX-CoV2373 having on average the shortest interval, the differences in disease risk between vaccine types were also observed in the booster dose analysis.

It is notable that the individuals receiving a primary course in this study period may have been vaccine hesitant given they had access to vaccination earlier in 2021; more than 91% of the adult Australian population was estimated to have received a primary course prior to 2022^13^. However, our study was robust because we only compared vaccine types between individuals receiving a primary course in the same calendar period. In Australia all vaccines were recommended for use in a primary course, but booster recommendations preferenced mRNA vaccines over other vaccine types^2^, therefore those who received NVX-CoV2373 or ChAdOx1 nCov-19 may have also had some contraindication to mRNA vaccine receipt. Whether this could independently alter their risk of SARS-CoV-2 infection is unclear. Also, in considering NVX-CoV2373 as a booster, as this vaccine was introduced much later, those who received it may be systematically different to the majority of the population who received the other 3 vaccine types in ways that we were unable to measure. Finally, we only compared the interval 2-9 weeks since vaccine receipt as too few individuals receiving NVX-CoV2373 had received vaccine >9 weeks earlier, given the timing of NVX-CoV2373 introduction into Australia. So, it is unclear if the differences we observed would continue to be seen with longer observation time following vaccine receipt.

In summary, our findings suggest relatively higher effectiveness of mRNA vaccines against SARS-CoV-2 Omicron infection than viral vector and protein subunit vaccines, when all vaccines were based on the ancestral strain. The findings are consistent with available immunological data, models predicting vaccine effectiveness by vaccine platform, and data from observational studies of mRNA and ChAdOx1 nCov-19, but provide important new information on NVX-CoV2373. From an epidemiological perspective, given the relatively small numbers of individuals receiving the viral vector and protein subunit vaccine, it would be useful to see these comparative effectiveness analyses repeated in other settings. From a public health perspective, we emphasise that as all four vaccines have been shown to provide good protection against severe disease from SARS-CoV-2 regardless of the variant type, our findings should not change the messaging around the importance of using available COVID-19 vaccines to protect populations at risk.

## Data Availability

All data produced in the present study are available upon reasonable request to the NSW Ministry of Health with appropriate ethical approvals.

## Acknowledgements

We thank colleagues at NSW Health for their support and access to data and to the NSW Centre for Health Record Linkage for conducting the linkage.

## Funding

Supported by NSW Ministry of Health

## Declarations

All authors have no conflicts of interest to declare

## Data Access

Data used in this analysis are accessible on application to the NSW Ministry of Health with appropriate ethical approvals.

---

**Supplementary Table 1:** Counts, rates and adjusted hazard ratios for SARS-CoV-2 infection by vaccine type in subgroups of age, gender and socioeconomic status

|  | Primary course; dose 2 14-63 days |  |  |  |  |  | Booster; dose 3 14-63 days |  |  |  |  |  |
| --- | --- | --- | --- | --- | --- | --- | --- | --- | --- | --- | --- | --- |
|  | Number of Infections | Person Years | Person-time rate (per 100-Person-Year) (95% CI) | Adjusted Hazard Ratio* | 95%CI |  | Number of Infections | Person Years | Person-time rate (per 100-Person-Year) (95% CI) | Adjusted Hazard Ratio* | 95%CI |  |
| **By age** |  |  |  |  |  |  |  |  |  |  |  |  |
| **18-49 years** |  |  |  |  |  |  |  |  |  |  |  |  |
| BNT162b2 | 490 | 1046 | 46.835 (39.112; 56.083) | 1.00 |  |  | 52742 | 61823 | 85.311 (83.908; 86.737) | 1.00 |  |  |
| mRNA-1273 | 68 | 135 | 50.524 (31.147; 81.956) | 1.07 | 0.83 | 1.38 | 14428 | 16303 | 88.497 (85.736; 91.347) | 1.03 | 1.01 | 1.05 |
| ChAdOx-1 nCov-19 | 64 | 103 | 62.092 (37.713; 102.231) | 1.35 | 1.04 | 1.75 | 227 | 217 | 104.664 (81.293; 134.754) | 1.28 | 1.13 | 1.46 |
| NVX-CoV2373 | 158 | 231 | 68.283 (49.716; 93.785) | 1.80 | 1.50 | 2.15 | 81 | 109 | 74.129 (48.559; 113.163) | 1.10 | 0.89 | 1.37 |
| **50+ years** |  |  |  |  |  |  |  |  |  |  |  |  |
| BNT162b2 | 145 | 484 | 29.982 (22.095; 40.684) | 1.00 |  |  | 20631 | 60908 | 33.872 (33.056; 34.709) | 1.00 |  |  |
| mRNA-1273 | 16 | 59 | 27.317 (10.898; 68.473) | 0.90 | 0.54 | 1.51 | 5089 | 14779 | 34.433 (32.782; 36.167) | 0.98 | 0.95 | 1.02 |
| ChAdOx-1 nCov-19 | 19 | 82 | 23.309 (10.030; 54.168) | 0.84 | 0.52 | 1.35 | 276 | 682 | 40.477 (32.778; 49.986) | 1.29 | 1.14 | 1.46 |
| NVX-CoV2373 | 73 | 179 | 40.817 (26.546; 62.760) | 1.35 | 1.02 | 1.79 | 106 | 162 | 65.517 (46.611; 92.091) | 1.70 | 1.41 | 2.31 |
| **By sex** |  |  |  |  |  |  |  |  |  |  |  |  |
| **Women** |  |  |  |  |  |  |  |  |  |  |  |  |
| BNT162b2 | 372 | 761 | 48.869 (39.902; 59.850) | 1.00 |  |  | 41529 | 64307 | 64.580 (63.416; 65.765) | 1.00 |  |  |
| mRNA-1273 | 50 | 88 | 56.683 (32.609; 98.531) | 1.14 | 0.85 | 1.53 | 10927 | 16264 | 67.187 (64.846; 69.612) | 1.01 | 0.99 | 0.99 |
| ChAdOx-1 nCov-19 | 40 | 86 | 46.647 (25.140; 86.553) | 1.08 | 0.78 | 1.50 | 258 | 428 | 60.270 (47.850; 75.914) | 1.28 | 0.78 | 1.13 |
| NVX-CoV2373 | 126 | 214 | 58.795 (41.503; 83.292) | 1.53 | 1.25 | 1.87 | 118 | 161 | 73.158 (52.008; 102.909) | 1.40 | 0.72 | 1.17 |
| **Men** |  |  |  |  |  |  |  |  |  |  |  |  |
| BNT162b2 | 263 | 769 | 34.217 (26.827; 43.644) | 1.00 |  |  | 31844 | 58425 | 54.504 (53.380; 55.652) | 1.00 |  |  |
| mRNA-1273 | 34 | 105 | 32.396 (16.465; 63.739) | 0.93 | 0.65 | 1.33 | 8590 | 14819 | 57.965 (55.686; 60.338) | 1.04 | 1.01 | 1.06 |
| ChAdOx-1 nCov-19 | 43 | 99 | 43.506 (23.834; 79.415) | 1.34 | 0.97 | 1.86 | 245 | 471 | 52.053 (41.047; 66.009) | 1.15 | 1.01 | 1.30 |
| NVX-CoV2373 | 105 | 196 | 53.589 (36.461; 78.764) | 1.94 | 1.55 | 2.43 | 69 | 110 | 62.862 (40.179; 98.349) | 1.37 | 1.08 | 1.73 |
| **By socioeconomic status** |  |  |  |  |  |  |  |  |  |  |  |  |
| **Lower** |  |  |  |  |  |  |  |  |  |  |  |  |
| BNT162b2 | 397 | 989 | 40.158 (32.982; 48.895) | 1.00 |  |  | 32865 | 62231 | 52.811 (51.741; 53.904) | 1.00 |  |  |
| mRNA-1273 | 59 | 128 | 46.099 (27.664; 76.818) | 1.12 | 0.86 | 1.48 | 8291 | 14838 | 55.876 (53.644; 58.201) | 1.04 | 1.01 | 1.06 |
| ChAdOx-1 nCov-19 | 45 | 110 | 41.019 (22.859; 73.608) | 1.12 | 0.83 | 1.53 | 250 | 502 | 49.842 (39.414; 63.028) | 1.22 | 1.07 | 1.38 |
| NVX-CoV2373 | 112 | 195 | 57.494 (39.688; 83.288) | 1.76 | 1.43 | 2.18 | 59 | 99 | 59.841 (36.911; 97.017) | 1.34 | 1.04 | 1.73 |
| **Higher** |  |  |  |  |  |  |  |  |  |  |  |  |
| BNT162b2 | 238 | 541 | 43.972 (34.085; 56.728) | 1.00 |  |  | 40508 | 60500 | 66.955 (65.732; 68.201) | 1.00 |  |  |
| mRNA-1273 | 25 | 65 | 38.358 (17.480; 84.171) | 0.88 | 0.58 | 1.32 | 11226 | 16245 | 69.106 (66.728; 71.569) | 1.00 | 0.98 | 1.02 |
| ChAdOx-1 nCov-19 | 38 | 75 | 50.745 (26.826; 95.991) | 1.26 | 0.89 | 1.77 | 253 | 397 | 63.702 (50.450; 80.435) | 1.21 | 1.07 | 1.37 |
| NVX-CoV2373 | 119 | 215 | 55.237 (38.530; 79.190) | 1.62 | 1.30 | 2.02 | 128 | 172 | 74.218 (53.469; 103.018) | 1.40 | 1.18 | 1.66 |

\*Adjusted for age, gender, socioeconomic status and comorbidity

**Supplementary Table 2:** Rates and adjusted hazard ratios for SARS-CoV-2 infection by vaccine type in all population (both those with and without previous infection)

|  | Number of Infections | Person Years | Person-time rate (per 100-Person-Year) (95% CI) | Adjusted Hazard Ratio* | 95%CI |  |
| --- | --- | --- | --- | --- | --- | --- |
| **Primary course; dose 2 14-63 days** |  |  |  |  |  |  |
| BNT162b2 (Pfizer Comirnaty) | 732 | 2224 | 32.917 (28.488; 38.033) | 1.00 |  |  |
| mRNA-1273 (Moderna Spikevax) | 88 | 235 | 37.442 (24.682; 56.798) | 0.96 | 0.77 | 1.20 |
| ChAdOx-1 nCov-19 | 97 | 253 | 38.362 (25.795; 57.053) | 1.22 | 0.99 | 1.51 |
| NVX-CoV2373 | 246 | 511 | 48.157 (37.533; 61.787) | 1.58 | 1.36 | 1.82 |
| **Booster; dose 3 14-63 days** |  |  |  |  |  |  |
| BNT162b2 (Pfizer Comirnaty) | 74049 | 142635 | 51.915 (51.212; 52.628) | 1.00 |  |  |
| mRNA-1273 (Moderna Spikevax) | 19694 | 36084 | 54.578 (53.154; 56.041) | 1.02 | 1.00 | 1.04 |
| ChAdOx-1 nCov-19 | 506 | 998 | 50.687 (42.978; 59.779) | 1.21 | 1.10 | 1.32 |
| NVX-CoV2373 | 191 | 318 | 60.008 (45.875; 78.493) | 1.39 | 1.20 | 1.60 |

\*Adjusted for age, gender, socioeconomic status, comorbidity, previous infection. Total study population included those with a prior COVID-19 diagnosis; in primary course analysis N=2,697,357; booster analysis N=5,248,353.

**Supplementary Table 3:** Analyses restricting study period to 1 April to 27 May 2022 for rates and adjusted hazard ratios for SARS-CoV-2 infection by vaccine type

|  | Number of Infections | Person Years | Person-time rate (per 100-Person-Year) (95% CI) | Adjusted Hazard Ratio* | 95%CI |  |
| --- | --- | --- | --- | --- | --- | --- |
| **Primary course; dose 2 14-63 days** |  |  |  |  |  |  |
| BNT162b2 (Pfizer Comirnaty) | 262 | 676 | 38.775 (29.075; 51.711) | 1.00 |  |  |
| mRNA-1273 (Moderna Spikevax) | 38 | 83 | 45.912 (21.558; 97.779) | 1.15 | 0.82 | 1.62 |
| ChAdOx-1 nCov-19 | 23 | 62 | 37.196 (14.076; 98.288) | 0.99 | 0.65 | 1.51 |
| NVX-CoV2373 | 216 | 393 | 54.968 (40.032; 75.478) | 1.71 | 1.43 | 2.05 |
| **Booster; dose 3 14-63 days** |  |  |  |  |  |  |
| BNT162b2 (Pfizer Comirnaty) | 20917 | 37441 | 55.867 (54.232; 57.551) | 1.00 |  |  |
| mRNA-1273 (Moderna Spikevax) | 5419 | 9460 | 57.286 (54.040; 60.728) | 1.01 | 0.98 | 1.04 |
| ChAdOx-1 nCov-19 | 194 | 322 | 60.173 (44.206; 81.908) | 1.32 | 1.15 | 1.52 |
| NVX-CoV2373 | 168 | 256 | 65.555 (47.064; 91.310) | 1.49 | 1.28 | 1.73 |

\*Adjusted for age, gender, socioeconomic status, and comorbidity

## Notes

### Competing Interest Statement

The authors have declared no competing interest.

### Funding Statement

This study was funded by the NSW Ministry of Health

### Author Declarations

The NSW Population and Health Services Research Ethics Committee gave ethical approval for this work.

### Summary of Updates

Author name correction, minor clarification to text.

